# Non-Linear Fitting of Sigmoidal Growth Curves to predict a maximum limit to the total number of COVID-19 cases in the United States

**DOI:** 10.1101/2020.04.22.20074898

**Authors:** Carlos Maximiliano Dutra

## Abstract

In the present work is used non-linear fitting of the “Gompert” and “Logistic” growth models to the number of total COVID-19 cases from the United States as a country and individually by states. The methodology allowed us to estimate that the maximum limit for the total number of cases of COVID-19 patients such as those registered with the World Health Organization will be approximately one million and one hundred thousand cases to the United States. Up to 04/19/20 the models indicate that United States reached 70% of this maximum number of “total cases” and the United States will reach 95% of this limit by 05/14/2020. The application of the nonlinear fitting of growth curves to the individual data of each American state showed that only 25% of them did not reach, on 04/19/20, the percentage of 59% of the maximum limit of “total cases” and that 17 of the 50 states still will not have reached 95% of that limit on 05/14/20.

## Introduction

The first case of SARS-CoV-2 (severe acute respiratory syndrome coronavirus 2) infection appeared in the Hubei Province in China on December 31, 2019, as reported by the World Health Organization - WHO (SR1, 2020). According to Chan et al. (2020) in february WHO officially called the disease caused by SARS-CoV-2 of COVID-19 (Coronavirus Dease 2019) and on 11 march the COVID-19 was classified as a Pandemic.

In the United States, the first case of COVID19 was registered on January 10, 2020 and by April 20, 2020, 792,759 total cases, +28,123 new cases and 42,514 deaths had been registered (https://www.worldometers.info/coronavirus/). For the United States according to the “…CDC case counts and death counts include boh confirmed and probable cases and deaths.” (CDC, 2000). Daily statistics on the number of new cases, total cases and deaths have been the main variables for controlling the impact of COVID19 on society and especially on the capacity of the hospital health system to take into account the number of critical cases. The real impact of COVID19 in the world will hardly be known as there are many asymptomatic cases and we have limitations in the ability to carry out tests to confirm this disease, although the United States was the country that performed the most tests 4,026,360 however it reached 12,164 per 1 million people, as data from April 20, 2020 (https://www.worldometers.info/coronavirus/country/us/).

Several researchers have presented models to predict the evolution of the number of cases of infected by COVID19. Bliznashki (2020) applied a Bayesian Logistic Growth model to the COVID-19 data cases in the period between 4^th^ of March and 31^th^ of March and found that the data in this interval have small number of available data points without upper asymptote to can be well fitted by this model, but it will be useful with more actualizated data. Mondal and Ghosh (2020) studied the scenarios of the exponential and sigmoid growth of COVID-19 total cases data for 15 states of India considering a initial exponential growth and a extrapolation with a sigmoid-type profile. Peirlinck et al. (2020) estimated to the United States the nationwide peak of the outbreak on May 10, 2020 with 3 million infections across the United States.

In the present work we propose the use of non-linear adjustment of sigmoidal growth functions together with the total case data to estimate maximum limits for the number of total cases of COVID19 in the United States.

## Metodology and Results

In order to predicted the maximum limit to the total cases COVID19 in the states of US was applyed to the data of cummulative cases a non-linear fitting of the Sigmoidal growth curves that are mathematical solutions to the infected cases growing models. According to Teleken et al. (2017):

> The growth functions can be grouped into three models main categories: those with a diminishing returns behavior (Brody model), those with sigmoidal shape and a fixed inflection point (Gompertz, Logistic and von Bertalanffy models) and those with a flexible inflection point (Richards model). The Logistic, Gompertz and von Bertalanffy models exhibit inflection points at about 50, 37 and 30% of the mature weight (asymptote), respectively. On the other hand, the Brody model does not exhibit an inflection point.

Sigmoid growth curves have been used to estimate the time evolution of total cases, daily cases and deaths for countries like Iran (Ahmadi et al. 2020) and China, Italy and Germany (Mimkes & Janssen 2020). This work will be carried out the non-linear fitting to the cummulative cases COVID-19 using Gompertz and Logistic models. The matemathical functions used were:

Gompertz Model:

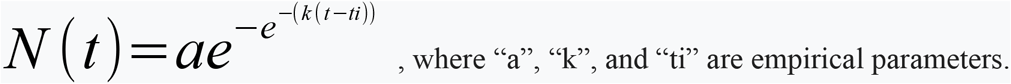

Logistic Model:

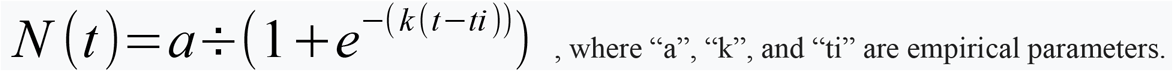

The parameter “a” gives the maximum limit to the total cases, the parameter “k” is related to the growth rate and “ti” corresponds to the inflection time coordinate.

The performance of each model was evaluated by Residual Sum Square (RSS) and two additional criteria based on the information theory were applied to compare the goodness of fit of the models (Burnham & Anderson, 2002): the Akaike information criterion (AIC) and the Bayesian information criterion (BIC).

## Results and discussion

First was performed non-linear fitting of Gompertz and Logistic models to the data of cummulative cases COVID19 of Hubei (China), South Korea and US, available in the site https://data.humdata.org/dataset/novel-coronavirus-2019-ncov-cases. and was obtain the results shown in Table 1. The Logistic Model give better goodness fit to the Hubei(China), while Gompertz model adjusted better the data from South Korea and United States. In sequence, Table 1 presents the fitting results for the states of the United States, which the data were obtained in the site https://github.com/nytimes/covid-19-data/blob/master/us-states.csv. Although for the US data the best fit model was the Gompertz, considering individual fits for 14 of the 50 american states the best fit model were the logistic. The data considered to fit were up to 04/17/20, and the best fit models have the parameters in bold.

**Table 1.** Results of Non-linear fitting from total cases COVID19 data up to 04/17/20.

|  | Gompertz Model |  |  |  |  |  | Logistic Model |  |  |  |  |  |
| --- | --- | --- | --- | --- | --- | --- | --- | --- | --- | --- | --- | --- |
| Country | a | xc | k | RSS | AIC | BIC | a | ti | k | RSS | AIC | BIC |
| Hubei, China | 68244,1 | 16,8 | 0,16 | 3,99E+008 | 1343,16 | 1352,54 | **67769,03** | **19,7** | **0,23** | **2,37E+008** | **1297,49** | **1306,86** |
| Korea South | **10125,85** | **40,13** | **0,13** | **1,01E+007** | **1023,3** | **1032,67** | 9854,84 | 43,05 | 0,2 | 1,95E+007 | 1080,21 | 1089,58 |
| US | **1122702,72** | **77,47** | **0,08** | **1,92E+008** | **1279,21** | **1288,59** | 782563,95 | 76,72 | 0,18 | 3,22E+009 | 1524,59 | 1533,97 |
| States of United States |  |  |  |  |  |  |  |  |  |  |  |  |
| State | a | xc | k | RSS | AIC | BIC | a | ti | k | RSS | AIC | BIC |
| Alabama | **11199,17** | **33,69** | **0,06** | **127491,34** | **303,49** | **308,54** | 5925,79 | 28,69 | 0,17 | 147477,47 | 308,73 | 313,78 |
| Alaska | **393,48** | **21,49** | **0,09** | **1060,13** | **133,39** | **138,59** | 323,55 | 23,01 | 0,17 | 1915,66 | 170,83 | 176,02 |
| Arizona | **6434,3** | **38,85** | **0,08** | **80404,14** | **378,03** | **384,79** | 4787,57 | 38,99 | 0,19 | 319241,13 | 446,97 | 453,73 |
| Arkansas | **3333,42** | **31,23** | **0,06** | **29156,96** | **261,64** | **266,98** | 2076,76 | 28,43 | 0,14 | 64365,31 | 291,73 | 297,07 |
| Califórnia | **45872,93** | **73,58** | **0,08** | **2299549,23** | **866,77** | **875,98** | 32541,15 | 73,05 | 0,17 | 4855789,13 | 929,55 | 938,77 |
| Colorado | **13395,26** | **32,39** | **0,08** | **294629,74** | **396,63** | **402,74** | 9861,07 | 32,4 | 0,17 | 1252439,33 | 460,31 | 466,42 |
| Connecticut | **34734,72** | **36,59** | **0,07** | **771814,01** | **412,67** | **418,41** | 20362,07 | 33,59 | 0,19 | 15578364,85 | 442 | 447,74 |
| Delaware | 6482,66 | 38,41 | 0,06 | 50986,99 | 282,88 | 288,22 | **2984,41** | **32,11** | **0,19** | **24517,76** | **255,05** | **260,39** |
| Columbia* | 5534,65 | 38,47 | 0,06 | 44338,19 | 301,48 | 307,35 | **3027,89** | **34,1** | **0,17** | **32426,87** | **288,34** | **294,21** |
| Florida | **33764,86** | **36,05** | **0,09** | **820812,24** | **476,78** | **483,33** | 26104,57 | 36,64 | 0,2 | 5220809,16 | 565,58 | 572,14 |
| Georgia | **28059,52** | **38,89** | **0,07** | **2874350,97** | **526,94** | **533,4** | 18634,22 | 37,48 | 0,18 | 3879116,33 | 541,04 | 547,49 |
| Guam* | 1160,87 | 24,78 | 0,12 | 52768,22 | 259,19 | 263,91 | **888,91** | **25,1** | **0,27** | **33610,01** | **243,85** | **248,58** |
| Hawaii | 654,88 | 26,17 | 0,1 | 2696,91 | 187,01 | 193,01 | **559,28** | **27,89** | **0,2** | **1526,09** | **162,53** | **168,52** |
| Idaho | 1594,27 | 18,89 | 0,19 | 58184,48 | 275,25 | 280,29 | **1500,56** | **20,6** | **0,32** | **53463,26** | **272,21** | **277,25** |
| Illinois | **57349,97** | **80,4** | **0,07** | **613843,53** | **763,71** | **772,98** | 33463,51 | 77,07 | 0,17 | 3898019,14 | 920,83 | 930,1 |
| Indiana | **14821,53** | **33,07** | **0,09** | **179034,27** | **367,42** | **373,41** | 10914,07 | 33,08 | 0,2 | 964801,91 | 439,85 | 445,84 |
| Iowa | **7672,64** | **44,88** | **0,05** | **13460,98** | **246,66** | **252,41** | 3254,77 | 35,83 | 0,15 | 44410,58 | 295,6 | 301,35 |
| Kansas | **2770,03** | **33,06** | **0,08** | **12566,67** | **248,53** | **254,4** | 1904,09 | 32,13 | 0,18 | 27775,92 | 281,84 | 287,71 |
| Kentucky | **5768,24** | **39,57** | **0,06** | **34454,74** | **296,56** | **302,55** | 3186,01 | 35,31 | 0,17 | 38643,21 | 301,49 | 307,48 |
| Louisiana | 26184,18 | 25,41 | 0,15 | 305406,99 | 511,2 | 516,81 | **23255,05** | **27,01** | **0,27** | **4386832,21** | **473,35** | **478,96** |
| Maine | **1240,53** | **24,96** | **0,07** | **8906,1** | **212,14** | **217,33** | 903,07 | 24,79 | 0,15 | 18660,84 | 239,51 | 244,7 |
| Maryland | 29483,08 | 42,91 | 0,07 | 704619,75 | 435 | 441,11 | **14766,82** | **37,8** | **0,19** | **481112,85** | **418,21** | **424,32** |
| Massachussets | **86495,04** | **75,77** | **0,06** | **3969635,04** | **844,03** | **852,85** | 44559,52 | 70,56 | 0,17 | 1,04E+007 | 918,21 | 927,03 |
| Michigan | **38301,45** | **26,06** | **0,11** | **2376608,87** | **438,86** | **444,34** | 31026,27 | 27,09 | 0,22 | 6424144,33 | 477,64 | 483,12 |
| Minnesota | **5831,74** | **44,3** | **0,04** | **27440,57** | **286,77** | **292,76** | 2774,94 | 36,1 | 0,13 | 67940,82 | 325,76 | 331,75 |
| Mississippi | **9580,03** | **36,85** | **0,05** | **60051,03** | **289,1** | **294,43** | 4949,3 | 31,01 | 0,15 | 194556,43 | 333,77 | 339,1 |
| Missouri | **7744,96** | **31,32** | **0,09** | **72905,86** | **322,37** | **328,24** | 5771,46 | 31,48 | 0,2 | 321060,88 | 384,63 | 390,5 |
| Montana | **463,77** | **17,64** | **0,12** | **986,28** | **128,46** | **133,51** | 418,9 | 19,75 | 0,22 | 2550,14 | 162,66 | 167,71 |
| Nebraska | 19949,46 | 97,46 | 0,03 | 10931,24 | 325,21 | 332,94 | **2285,38** | **60,46** | **0,14** | **7759,51** | **304,31** | **312,04** |
| Nevada | **5039,34** | **31,93** | **0,08** | **30529,53** | **296,88** | **302,99** | 3791,19 | 32,23 | 0,18 | 144881,65 | 365,4 | 371,51 |
| New Hampshire | **1971,13** | **36,78** | **0,07** | **14543,43** | **278,48** | **284,93** | 1391,82 | 36,19 | 0,17 | 28280,37 | 309,74 | 316,19 |
| New Jersey | **113272,31** | **34,34** | **0,09** | **9289561,43** | **559,7** | **565,92** | 84799,07 | 34,55 | 0,2 | 7,24E+007 | 652,09 | 658,32 |
| New Mexico | 4531,08 | 37,55 | 0,06 | 12127,37 | 228,31 | 233,64 | **2199,88** | **31,36** | **0,17** | **7631,16** | **210,7** | **216,04** |
| New York | **340941,99** | **36,3** | **0,08** | **1,02E+008** | **708,39** | **714,94** | 251484,83 | 36,34 | 0,17 | 7,64E+008 | 804,91 | 811,46 |
| North Carolina | **10439,31** | **38,59** | **0,07** | **83665,4** | **354,24** | **360,59** | 6731 | 36,71 | 0,17 | 281144,65 | 410 | 416,34 |
| North Dakota | **782,81** | **31,08** | **0,06** | **2687,29** | **171,04** | **176,38** | 493,62 | 28,63 | 0,15 | 5316,08 | 196,97 | 202,3 |
| Ohio | **15785,09** | **32,66** | **0,07** | **460996,81** | **383,23** | **388,84** | 10270,33 | 30,91 | **0,17** | 1442560,39 | 428,86 | 434,48 |
| Oklahoma | **3317,6** | **31,1** | **0,1** | **26953,48** | **286** | **291,99** | 2595,19 | 31,79 | 0,21 | 72332,65 | 328,45 | 334,44 |
| Oregon | **2430,03** | **35,96** | **0,08** | **11854,37** | **282,31** | **289,07** | 1901,56 | 36,78 | 0,17 | 28816,92 | 326,72 | 333,48 |
| Pennsylvania | **33453,22** | **34,15** | **0,21** | **2963144,25** | **476,49** | **482,48** | 33453,22 | 34,15 | 0,21 | 2963144,25 | 488,1 | 494,09 |
| **Puerto Rico\*** | **1553,21** | **25,81** | **0,1** | **7616,16** | **202,05** | **228,59** | 1174,75 | 26,13 | 0,21 | 15918,41 | 228,59 | 233,64 |
| Rhode Island | 36532,64 | 65,81 | 0,04 | 113132,08 | 381,65 | 388,21 | **6957,21** | **45,78** | **0,19** | **55548,44** | **347,51** | **354,06** |
| Sourh Carolina | **5844,49** | **31,56** | **0,09** | **84398,48** | **335,08** | **341,07** | 4378,93 | 31,77 | 0,19 | 129010,5 | 353,33 | 359,32 |
| South Dakota | 21496 | 64,37 | 0,04 | 28939 | 223,43 | 228,91 | **2889,7** | **39,33** | **0,19** | **13652** | **209,22** | **214,69** |
| Tennessee | **8599,3** | **30,79** | **0,09** | **243574,75** | **388,26** | **394,37** | 6700,53 | 31,53 | 0,18 | 737350,93 | 437 | 443,11 |
| Texas | 33120,41 | 59,38 | 0,07 | 1274303,81 | 659,96 | 668,06 | **20979,67** | **57,39** | **0,19** | **1,26E+006** | **659,27** | **667,37** |
| Utah | **3842,86** | **40,19** | **0,08** | **37097,35** | **356,04** | **363,08** | 2945,26 | 40,7 | 0,18 | 8,26E+004 | 398,48 | 405,53 |
| Vermont | 989,04 | 26,46 | 0,1 | 10151,68 | 239,56 | 245,43 | **831,15** | **28,02** | **0,2** | **6,51E+003** | **220,9** | **226,77** |
| Virginia | **18998,42** | **40,96** | **0,06** | **120044,57** | **343,31** | **349,18** | 9587,79 | 35,61 | 0,18 | 2,01E+005 | 365,06 | 370,93 |
| Washington | 14714,5 | 70,55 | 0,08 | 1948290,21 | 888,93 | 898,36 | **12023,21** | **72,02** | **0,17** | **724686,95** | **801,9** | **811,33** |
| West Virginia | 1297,2 | 23,59 | 0,08 | 6419,71 | 179,13 | 183,51 | **885,66** | **22,64** | **0,2** | **5223,5** | **172,53** | **176,91** |
| Wisconsin | **5763,44** | **60,24** | **0,08** | **34957,12** | **459,1** | **467,67** | 4325,38 | 60,51 | 0,18 | 181686,31 | 579,42 | 587,99 |
| Wyoming | 369,85 | 21,85 | 0,1 | 1164,56 | 139,27 | 144,61 | **311,66** | **23,42** | **0,2** | **630,95** | **115,98** | **121,32** |

\* In addition to the 50 states, two territories Guam and Puerto Rico and the district of Columbia were included.

Defined the best model to adjust the total case data, the model predictions were made using the parameters “a” to define the maximum of the total cases and the other empirical parameters and the mathematical functions to make temporal predictions for the data observed on 4/19/20 and the determination of the dates on which 95% and 99% of the maximum total cases will be reached, according to Table 2. Model forecasts were made for Hubei (China) and South Korea, which are already in the stable control phase of COVID19 and the results of the total number of cases in the model adjusted well to the data of total cases observed on April 19, 2020 (comparing the predicted cases with those observed and verifying the low relative error value). The maximum percentage reached according to the model for that date was also estimated, as well as the forecast of dates when according to the model a percentage of 95% and 99%, respectively, of the maximum value of the total cases predicted in the model are reached. For the total case data from the United States, we had the Gompertz model as the best fit, from which we have an estimated maximum for the total MAX_Cases cases of 1,122,702.72. For April 19, 2020, the comparison between the number of cases predicted by the model and those observed results in a relative error of −0.62% and on this day the total number of cases predicted by the model corresponds to a percentage of 67, 19% in relation to the maximum of MAX_Cases total cases. Finally, the model predicts that the United States will reach 95% of the maximum total number of cases on 5/14/20 and 99% on 6/4/20.

**Table 2.** Predictions for the “total cases” data.

| Country | Model | MAX_Cases | Predictions 04/19/20 |  |  |  | Date 95% | Date 99% |
| --- | --- | --- | --- | --- | --- | --- | --- | --- |
|  |  |  | Predicted Cases | Observed Cases | Relative Error(%) | Pr_max (%) |  |  |
| Hubei, China | L | 67769,03 | 67769,02 | 68128 | -0,53 | 99,99 | 02/22/20 | 02/29/20 |
| Korea South | G | 10125,85 | 10108,23 | 10661 | -5,18 | 99,83 | 03/24/20 | 04/05/20 |
| US | G | 1122702,72 | 754405,83 | 759086 | -0,62 | 67,19 | 05/14/20 | 06/04/20 |

|  |  |  | Predictions 04/19/20 |  |  |  |  |  |
| --- | --- | --- | --- | --- | --- | --- | --- | --- |
| State | Model | MAX_Cases | Predicted Cases | Observed Cases | Relative Error(%) | Pr_max (%) | Date 95% | Date 99% |
| Alabama | G | 11199,17 | 5174,32 | 4903 | 5,53 | 46,2 | 06/03/20 | 06/30/20 |
| Alaska | G | 393,48 | 319,96 | 317 | 0,93 | 81,32 | 05/04/20 | 05/22/20 |
| Arizona | G | 6434,3 | 4537,63 | 4929 | -7,94 | 70,52 | 05/13/20 | 06/06/20 |
| Arkansas | G | 3333,42 | 1846,24 | 1781 | 3,66 | 55,39 | 05/30/20 | 06/25/20 |
| Califórnia | G | 45872,93 | 31678,36 | 31544 | 0,43 | 69,06 | 05/14/20 | 06/03/20 |
| Colorado | G | 13395,26 | 9566,63 | 9730 | -1,68 | 71,42 | 05/12/20 | 02/01/20 |
| Connecticut | G | 34734,72 | 18343,63 | 17962 | 2,12 | 52,81 | 05/25/20 | 06/17/20 |
| Delaware | L | 2984,41 | 2439,58 | 2538 | -3,87 | 81,74 | 04/26/20 | 05/05/20 |
| Columbia | L | 3027,89 | 2553,42 | 2793 | -8,58 | 84,33 | 04/26/20 | 05/06/20 |
| Florida | G | 33764,86 | 25393,35 | 26306 | -3,47 | 75,21 | 05/08/20 | 05/26/20 |
| Georgia | G | 28059,52 | 17142,34 | 17619 | -2,7 | 61,09 | 05/21/20 | 06/13/20 |
| Guam | L | 888,91 | 844,4 | 808 | 4,51 | 94,99 | 04/19/20 | 04/25/20 |
| Hawaii | L | 559,28 | 541,6 | 574 | -5,64 | 96,84 | 04/16/20 | 04/24/20 |
| Idaho | L | 1500,56 | 1494,85 | 1581 | -5,44 | 99,62 | 04/10/20 | 04/15/20 |
| Illinois | G | 57349,97 | 30543,44 | 30357 | 0,61 | 53,26 | 05/25/20 | 06/17/20 |
| Indiana | G | 14821,53 | 10531,17 | 11210 | -6,06 | 71,05 | 05/10/20 | 05/28/20 |
| Iowa | G | 7672,64 | 2557,68 | 2915 | -12,26 | 33,33 | 06/19/20 | 07/21/20 |
| Kansas | G | 2770,03 | 1825,91 | 1898 | -3,8 | 65,92 | 05/15/20 | 06/04/20 |
| Kentucky | G | 5768,24 | 2802,23 | 2960 | -5,32 | 48,58 | 06/02/20 | 06/29/20 |
| Louisiana | L | 23255,05 | 22855,77 | 23928 | -4,48 | 98,28 | 04/15/20 | 04/21/20 |
| Maine | G | 1240,53 | 853,23 | 867 | -1,59 | 68,77 | 05/17/20 | 06/09/20 |
| Maryland | L | 14766,82 | 12198,37 | 12830 | -4,92 | 82,61 | 04/26/20 | 05/05/20 |
| Massachussets | G | 86495,04 | 37949,81 | 38077 | -0,33 | 43,87 | 06/04/20 | 07/01/20 |
| Michigan | G | 38301,45 | 31568,7 | 31348 | 0,7 | 82,42 | 05/01/20 | 05/15/20 |
| Minnesota | G | 5831,74 | 2205,44 | 2356 | -6,39 | 37,82 | 07/02/20 | 08/11/20 |
| Mississippi | G | 9580,03 | 4077,17 | 4274 | -4,6 | 42,55 | 06/14/20 | 07/16/20 |
| Missouri | G | 7744,96 | 5627,17 | 5667 | -0,7 | 72,66 | 05/09/20 | 05/27/20 |
| Montana | G | 463,77 | 425,17 | 433 | -1,81 | 91,68 | 04/23/20 | 05/07/20 |
| Nebraska | L | 2285,38 | 1343,75 | 1474 | -8,84 | 58,8 | 05/07/20 | 05/19/20 |
| Nevada | G | 5039,34 | 3643,04 | 3741 | -2,62 | 72,29 | 05/12/20 | 06/01/20 |
| New Hampshire | G | 1971,13 | 1288,52 | 1392 | -7,43 | 65,37 | 05/19/20 | 06/11/20 |
| New Jersey | G | 113272,31 | 74064,52 | 85301 | -13,17 | 65,39 | 05/15/20 | 06/04/20 |
| New Mexico | L | 4531,08 | 1788,23 | 1845 | -3,08 | 81,29 | 04/27/20 | 05/07/20 |
| New York | G | 340941,99 | 244082,7 | 242817 | 0,52 | 71,59 | 05/12/20 | 06/01/20 |
| North Carolina | G | 10439,31 | 6221,77 | 6493 | -4,18 | 59,6 | 05/22/20 | 06/14/20 |
| North Dakota | G | 782,81 | 435,87 | 585 | -25,49 | 55,68 | 05/30/20 | 06/25/20 |
| Ohio | G | 15785,09 | 9383,94 | 11602 | -19,12 | 59,45 | 05/22/20 | 06/14/20 |
| Oklahoma | G | 3317,6 | 2586,14 | 2599 | -0,49 | 77,95 | 05/05/20 | 05/21/20 |
| Oregon | G | 2430,03 | 1841,82 | 1910 | -3,57 | 75,79 | 05/10/20 | 05/30/20 |
| Pennsylvania | G | 33453,22 | 30196,02 | 32992 | -8,47 | 90,26 | 04/22/20 | 04/30/20 |
| Puerto Rico | G | 1553,21 | 1155,81 | 1213 | -4,71 | 74,41 | 05/06/20 | 05/22/20 |
| Rhode Island | G | 6957,21 | 4442,68 | 4706 | -5,59 | 63,86 | 04/30/20 | 05/09/20 |
| South Carolina | G | 5844,49 | 4337 | 4377 | -0,91 | 74,21 | 05/08/20 | 05/26/20 |
| South Dakota | L | 2889,7 | 1672,17 | 1635 | 2,27 | 57,86 | 05/03/20 | 05/11/20 |
| Tennessee | G | 8599,3 | 6667,83 | 6845 | -2,59 | 77,54 | 05/07/20 | 05/24/20 |
| Texas | L | 20979,67 | 18513,64 | 19443 | -4,77 | 88,24 | 04/24/20 | 05/02/20 |
| Utah | G | 3842,86 | 2830,37 | 3071 | -7,83 | 73,65 | 05/11/20 | 05/31/20 |
| Vermont | L | 831,15 | 798,47 | 812 | -1,67 | 96,07 | 04/17/20 | 04/26/20 |
| Virginia | G | 18998,42 | 8257,21 | 8537 | -3,28 | 43,46 | 06/04/20 | 07/01/20 |
| Washington | L | 12023,21 | 11482,97 | 11805 | -2,73 | 95,51 | 04/18/20 | 04/28/20 |
| West Virginia | L | 885,66 | 802,88 | 890 | -9,79 | 90,65 | 04/22/20 | 04/30/20 |
| Wisconsin | G | 5763,44 | 4239,74 | 4346 | -2,44 | 73,56 | 05/11/20 | 05/31/20 |
| Wyoming | L | 311,66 | 300,74 | 313 | -3,92 | 96,5 | 04/17/20 | 04/25/20 |
| TOTAL |  | 1085939,78 |  |  |  |  |  |  |

Note: in the "model" column letter "L" designate Logistic and letter "G" designate Gompertz.

Considering the COVID19 total case data from the states (plus Columbia, Guam and Puerto Rico) in the United States, the following results were summarized in Table 2: (a) limit for the maximum total cases for the United States of 1,085,939.78, which is approximately 3% less than the predicted total considering the data for the United States as a whole; (b) Only 25% of the states had a relative error greater than 6%, the average relative error in module was 4.90% for the value of “total cases” on 04/19/20; (c) Only 25% of the states reached less than 59% of the maximum value of “total cases” on 04/19/20; (d) Until 05/14/2020, only 17 of the 50 American states will not have reached 95% of the maximum limit of the “total cases”.

## Final considerations

The non-linear fitting of growth functions to the data on the total number of COVID-19 cases is a useful tool to estimate the duration and impact of this pandemic, and may become increasingly accurate as the history of the disease evolves. This study demonstrates the importance of carrying out the COVID-19 tests for understanding the future of the development of this pandemic, only with precise and actualizated statistics will be possible to prepare the health system to the demand.

## Data Availability

The data are all available.

